# Job stress and loneliness among remote workers

**DOI:** 10.1101/2021.05.31.21258062

**Authors:** Fuyu Miyake, Chimed-Ochir Odgerel, Ayako Hino, Kazunori Ikegami, Tomohisa Nagata, Seiichiro Tateishi, Mayumi Tsuji, Shinya Matsuda, Tomohiro Ishimaru

## Abstract

**Background:** To prevent the spread of coronavirus disease 2019 (COVID-19), physical distancing and isolation are crucial strategies in society. However, this response to the pandemic promotes loneliness. Previous studies have reported an increase in loneliness since the outbreak of COVID-19, but there is little evidence on the relationship between job stress and loneliness among remote workers.

**Aims:** To assess the relationship between job stress and loneliness among remote workers.

**Methods:** This study is a part of nation-wide cross-sectional online survey evaluating the impact of the COVID-19 pandemic in Japan. A total of 27,036 full-time workers completed the self-administrated questionnaire in December 2020. We extracted data on 4,052 desk workers who indicated that they were doing remote work. Loneliness was assessed using a single question and job stress was measured using the Job Content Questionnaire. Multiple logistic regression was performed.

**Results:** Frequency of remote work was moderately associated with loneliness (adjusted odds ratio [AOR] = 1.60, 95% confidence interval [CI]: 1.04–2.46, *P* = 0.033). Participants who reported of having a low level of co-worker or supervisor support had greater odds of feeling lonely than those who were highly supported (co-worker support: AOR = 4.06, 95% CI: 2.82–5.84, *P* <0.001; supervisor support: AOR = 2.49, 95% CI: 1.79–3.47, *P* <0.001).

**Conclusions:** Co-worker support and supervisor support were strongly associated with loneliness, whereas frequency of remote work was moderately associated with feeling lonely. Support from co-workers and supervisors may be crucial factors to prevent loneliness caused by remote work.

## Introduction

Loneliness, which has recently become a global concern, is generally defined as a discrepancy between an individual’s preferred and actual levels of social relations [1]. This discrepancy leads to anxiety and distress because of the negative experience of feeling alone [2]. Loneliness relates not only to stressful and unpleasant feelings but also to critical physical and psychological health issues. Although loneliness has often been regarded as a problem affecting older adults, it is also a risk for younger people [3]. According to a recent study on adults with loneliness [4], the prevalence of loneliness among adults aged 19 to 65 years was around 40% to 48%, showing that loneliness is a critical issue for the working generation. Although many factors are related to loneliness among adults, the major factors are considered to be socioeconomic status and income [5]. In addition, high population density is a robust correlate of loneliness [6]. Living alone and frequency of communication with neighbours have also been shown to be associated with loneliness [7]. With the addition of coronavirus disease 2019 (COVID-19), even more concern has been raised by the social issues of loneliness and its related.

Since the outbreak of the COVID-19 pandemic, work styles have changed dramatically, especially for desk workers. In February 2020, the Japanese government issued a basic policy describing measures against COVID-19. These measures included a recommendation that companies implement teleworking to prevent the spread of COVID-19 [7]. Following the continued spread of the disease throughout Japan, the government declared a state of emergency in April 2020, further promoting telework and requesting that people refrain from going out [8]. As a result, in Japan, the percentage of companies implementing telework climbed from 26% in March 2020 to 67% May 2020 [9]. Even after the state of emergency ended, an increasing number of companies continued to implement anti-COVID-19 measures by combining in-person work with remote work [10]. However, the impact of job stress on remote workers is unknown, especially in terms of loneliness. Previous research about teleworking in other countries has indicated that remote workers tend not to be able to establish social work relationships with others and that telework can induce feelings of loneliness [11]. It has been suggested that office workers should spend at least 20% of their work time in the office to prevent feelings of isolation [12].

Loneliness was already a critical issue before COVID-19. Physical distancing was then introduced as a crucial societal strategy to prevent the spread of COVID-19; this response to the pandemic further promotes loneliness. Several studies have revealed an increase in loneliness since the outbreak of COVID-19 [13], but no reports have clarified how job stress influences loneliness among remote workers during the pandemic. Therefore, the purpose of this study was to assess the relationship between job stress and loneliness among remote workers. The results will be useful for interventions targeted towards remote workers experiencing loneliness to improve the situation in the work environment.

## Methods

We conducted a cross-sectional study about COVID-19 among the working-age population in Japan on December 22–26, 2020, under the CORoNaWork (Collaborative Online Research on the Novel-coronavirus and Work) Project [14]. In brief, the CORoNaWork Project is an Internet-based nationwide cross-sectional study conducted during the third wave of COVID-19 infections in Japan. A total of 33,087 participants were selected using cluster sampling with stratification by sex, job type, and region on the basis of COVID-19 incidence rate data. These participants completed an online self-administered questionnaire. After excluding invalid responses, 27,036 participants were eligible for the analysis. In the current study, from these 27,036 participants, we selected the 4,052 desk workers who indicated that they were doing remote work. The study was conducted according to the guidelines of the Declaration of Helsinki, and it was approved by the Ethics Committee of the University of Occupational and Environmental Health, Japan (Approval number: R2-079).

The original questionnaire of the CORoNaWork Project consists of 54 main questions including items on general demographic characteristics, socioeconomic characteristics, work-related characteristics, lifestyle factors, quality of life, health conditions, and COVID-19-related issues (e.g. preventive measures at the workplace and individual levels, vaccination, telework, and lifestyle changes during COVID-19). For the current study, we included questions on demographic characteristics (age and sex), socioeconomic characteristics (education, annual household income, and household composition), regional state-of-emergency status, frequency of remote work, job stress, and loneliness.

We asked the participants whether they felt lonely during the study period. Loneliness was assessed by a single question: ‘Do you feel alone?’ The response options were *yes* and *no*. The question is a part of the Japanese version of the University of California, Los Angeles (UCLA) Loneliness Scale [15].

We included frequency of remote work and job stress as independent variables in the current study. Frequency of remote work was categorized as once per week, 2 or 3 days per week, or 4 or more days per week. We used the Job Content Questionnaire (JCQ) to evaluate the remote workers’ job stress. The JCQ is a self-administered instrument proposed by Karasek in 1985 [16] that was designed to measure the social and psychological characteristics of jobs on the basis of theoretical models. The original instrument comprises 45 core items; however, in 1995, Japanese researchers first translated the JCQ and developed the Japanese version of the JCQ with 22 items on four topics: co-worker support, supervisor support, psychological job demands, and decision latitude. The researchers then verified the reliability and validity of the questionnaire among the employees of telecommunication and electric power companies in the Chubu region [17] and among the workers of a computer company [18]. The average score and reliability coefficient of the Japanese version of the JCQ are very similar to the results in other countries; thus, the JCQ is considered to be internationally applicable in occupational settings [16]. Therefore, we used the Japanese version of the JCQ to assess job stress in the current research.

We present descriptive statistics of demographic and socioeconomic characteristics for all participants. Each of the 22 JCQ items had a four-point response ranging from 1 (*strongly disagree*) to 4 (*strongly agree*). The weighted item scores were summed to produce scores on the following four scales, following the authors of the Japanese version of the JCQ: the psychological demands scale (five items, range: 12–48), the decision latitude scale (nine items, range: 24–96), the co-worker support scale (four items, range: 4–16), and the supervisor support scale (four items, range: 4–16) [17]. Each sub-scale was classified into tertiles based on the sample distributions. The *high* group was used as a reference group for co-worker support, supervisor support, and decision latitude, whereas the *low* group was taken as a reference for psychological job demands. Logistic regression analysis was performed to identify the associations of frequency of remote work and the four JCQ scale scores with loneliness. We show the results of both the univariate model and the model adjusting for sex, age, education, income, household composition, and regional state-of-emergency status. Statistical significance was defined as *P* <0.05. Stata/SE□16.1 (StataCorp, College Station, TX, USA) was used for the statistical analysis.

## Results

A total of 4,052 remote workers were analysed in the current study. Table 1 shows the socio-demographic characteristics of the participants. Over half of the respondents were male (58%), were aged 50–65 years (52%), and had completed a university or graduate school degree (67%). Of the 4,052 remote workers participating in the study, 2,042 (50%) worked remotely 4 or more days per week, 1,058 (26%) worked remotely 2 or 3 days per week, and 952 (24%) worked remotely 1 day per week. Regarding job stress, almost half of all remote workers felt a high level of support from their co-workers (46%) and supervisors (49%). A total of 191 (5%) workers reported feeling lonely.

**Table 1.** General characteristics of the study participants

|  | Total<br>*n* = 4,052 |
| --- | --- |
| Sex |  |
| Female | 1,689 (42) |
| Male | 2,363 (58) |
| Age (years) |  |
| 20–39 | 807 (20) |
| 40–49 | 1,128 (28) |
| 50–65 | 2,117 (52) |
| Education |  |
| Junior high or high school | 617 (15) |
| Vocational school or college | 734 (18) |
| University or graduate school | 2,701 (67) |
| Annual household income (Japanese yen) |  |
| < 4 million | 890 (22) |
| ≥ 4 million and < 8 million | 1,554 (38) |
| ≥ 8,000,000 | 1,608 (40) |
| Household composition |  |
| Single | 912 (22) |
| Couple | 1,156 (29) |
| 3 or more persons | 1,984 (49) |
| Regional state-of-emergency status |  |
| No (34 prefectures) | 1,485 (37) |
| Yes (13 prefectures) | 2,567 (63) |
| Co-worker support |  |
| High (12–16 points) | 1,877 (46) |
| Middle (10 or 11 points) | 1,170 (29) |
| Low (4–9 points) | 1,005 (25) |
| Supervisor support |  |
| High (12–16 points) | 1,965 (49) |
| Middle (9–11 points) | 609 (15) |
| Low (4–8 points) | 1,478 (36) |
| Psychological job demand |  |
| High (32–48 points) | 1,362 (34) |
| Middle (27–31 points) | 1,341 (33) |
| Low (12–26 points) | 1,349 (33) |
| Decision latitude |  |
| High (71–96 points) | 1,262 (31) |
| Middle (63–70 points) | 1,628 (40) |
| Low (26–62 points) | 1,162 (29) |
| Frequency of remote work |  |
| 1 day/week | 952 (24) |
| 2 or 3 days/week | 1,058 (26) |
| 4 or more days/week | 2,042 (50) |
| Loneliness |  |
| Yes | 191 (5) |
| No | 3,861 (95) |

Table 2 shows the association between job stress and loneliness among remote workers. The highest percentage of loneliness was observed in the group with a low level of support from their co-workers (11%). In the multivariate model, participants who worked remotely 4 or more days per week had significantly greater odds of feeling lonely than those who worked at home once per week (adjusted odds ratio [AOR] = 1.60, 95% confidence interval [CI]: 1.04–2.46, *P* = 0.033). Participants who reported having a low level of co-worker support had greater odds of feeling lonely than those who were highly supported by their co-workers (AOR = 4.06, 95% CI: 2.82–5.84, *P* <0.001). Those who were less supported by their supervisors also had greater odds of feeling lonely than those who were highly supported by their supervisors (AOR = 2.49, 95% CI: 1.79–3.47, *P* <0.001). Compared with those who had low psychological job demands, participants who had high demands felt more loneliness (AOR = 2.04, 95% CI: 1.39–2.99, *P* <0.001). No significant associations were found between decision latitude and feeling lonely.

**Table 2.** Association between job stress and loneliness among remote workers (n=4,052)

|  | Participants | Loneliness | Univariate |  |  | Adjusted* |  |  |
| --- | --- | --- | --- | --- | --- | --- | --- | --- |
|  | *n* | % | OR | (95% CI) | *P* value | OR | (95% CI) | *P* value |
| Frequency of remote work |  |  |  |  |  |  |  |  |
| 1 day/week | 952 | 3 | 1.00 | - | - | 1.00 | - | - |
| 2 or 3 days/week | 1,058 | 5 | 1.60 | (0.99–2.57) | 0.049 | 1.59 | (0.98–2.56) | 0.059 |
| 4 or more days/week | 2,042 | 6 | 1.95 | (1.28–2.97) | 0.002 | 1.60 | (1.04–2.46) | 0.033 |
| Co-worker support |  |  |  |  |  |  |  |  |
| High (12–16 points) | 1,877 | 3 | 1.00 | - | - | 1.00 | - | - |
| Middle (10 or 11 points) | 1,170 | 3 | 1.34 | (0.86–2.07) | 0.191 | 1.33 | (0.85–2.06) | 0.209 |
| Low (4–9 points) | 1,005 | 11 | 4.74 | (3.33–6.76) | <0.001 | 4.06 | (2.82–5.84) | <0.001 |
| Supervisor support |  |  |  |  |  |  |  |  |
| High (12–16 points) | 1,965 | 3 | 1.00 | - | - | 1.00 | - | - |
| Middle (9–11 points) | 609 | 3 | 1.02 | (0.60–1.75) | 0.944 | 1.05 | (0.61–1.80) | 0.872 |
| Low (4–8 points) | 1,478 | 8 | 2.85 | (2.06–3.94) | <0.001 | 2.49 | (1.79–3.47) | <0.001 |
| Psychological job demand |  |  |  |  |  |  |  |  |
| High (32–48 points) | 1,362 | 5 | 1.71 | (1.18–2.47) | 0.004 | 2.04 | (1.39–2.99) | <0.001 |
| Middle (27–31 points) | 1,341 | 4 | 1.06 | (0.73–1.55) | 0.752 | 1.10 | (0.75–1.62) | 0.619 |
| Low (12–26 points) | 1,349 | 5 | 1.00 | - | - | 1.00 | - | - |
| Decision latitude |  |  |  |  |  |  |  |  |
| High (71–96 points) | 1,262 | 4 | 1.00 | - | - | 1.00 | - | - |
| Middle (63–70 points) | 1,628 | 4 | 0.81 | (0.56–1.17) | 0.258 | 0.80 | (0.55–1.17) | 0.250 |
| Low (26–62 points) | 1,162 | 7 | 1.11 | (0.79–1.56) | 0.560 | 0.99 | (0.69–1.42) | 0.945 |

OR: odds ratio; CI: confidence interval
\* Adjusted for sex, age, education, annual household income, household composition, and regional state-of-emergency status.

## Discussion

The present study revealed that frequency of remote work was moderately associated with loneliness among remote workers in Japan. In addition, co-worker and supervisor support and psychological job demands were related to loneliness, whereas decision latitude was not. These findings provide insight into strategies for improving loneliness resulting from working remotely during the COVID-19 pandemic.

Previous studies in other countries have reported that telework is associated with isolation and loneliness [11, 19]; we found a moderate relationship between frequency of remote work and feeling lonely among Japanese workers in our study. Generally, remote workers are thought to be separated from their colleagues and from work-related social relationships. Consequently, remote workers tend to have fewer opportunities for work-related social interaction and are also distanced from the specific instructions, attention, and praise of their supervisors. Being physically distant from the workplace and from one’s colleagues can lead to feelings of isolation and loneliness. This is regarded as the principal problem with telework [19]. Another study found that informal social interaction with colleagues declines for employees working at home, leading to professional isolation [20]. However, in recent years, advanced information and communication technology (ICT) has provided remote workers with opportunities for real - time social interaction [21], which may keep people socially connected and help to overcome feelings of loneliness [22]. Our study was conducted in December 2020, and we assume that most of the remote worker participants had the necessary ICT to work from home. In this situation, remote workers’ loneliness might be reduced by using the ICT that has been developed for this purpose. In this study, the frequency of telecommuting was found to be moderately related to feeling lonely. However, a previous study showed that the number of teleworking days per week was not associated with work-related well-being [23], so it may be important to consider how telecommuting is implemented to prevent loneliness and improve workers’ well-being.

Our analysis showed that the levels of support provided by co-workers and supervisors were strongly associated with feelings of loneliness among remote workers. Co-worker support contributed more to reducing loneliness than did supervisor support. A previous study indicated that the subjective experience of feeling physically distant from one’s colleagues increased loneliness and was stressful for remote workers [24], which can be taken to suggest the importance of support and connection among colleagues. Furthermore, although there are many opportunities to interact with supervisors through work instructions, even in the context of remote work, the frequency of communication between co-workers may be lower in remote work situations unless the co-workers make a conscious effort to stay in touch. Support from colleagues is likely to be particularly important in preventing the reduction of communication between co-workers.

The present study suggests that, although a middle level of psychological job demands did not affect the presence of loneliness, a high level of psychological job demands was associated with loneliness among remote workers. The presence of loneliness was also not affected by decision latitude. According to Karasek’s Demand–Control Model, a higher level of psychological job demands and a lower level of decision latitude result in an employee experiencing a high level of strain [25]; however, we found that, in terms of loneliness, only a high level of psychological job demands was related to loneliness. Therefore, our study indicates that job stress factors are not always associated with loneliness. Previous research has suggested that, if workers face higher levels of psychological work demands, they are more likely to work overtime [26]. As a result, one would expect those in jobs with high psychological demands to decrease the time spent communicating with colleagues and supervisors, which might lead to the experience of loneliness. Regarding decision latitude, it is generally thought that those with lower levels of decision latitude are managed or instructed by their supervisors and senior colleagues. To some extent, receiving instructions from others, as a form of communication, may play a role in preventing these workers from feeling lonely, although a low level of decision latitude is also a known stress factor for workers [25].

To the best of our knowledge, the present study is the first to report an association between job stress and loneliness among remote workers in Japan during the COVID-19 pandemic. However, there are several limitations in our study. First, the present study was conducted over the Internet, and the generalization of the results is thus unclear. However, to increase the external validity and reduce bias as much as possible, we defined the target population according to sex, job type, and region on the basis of COVID-19 incidence rate data. In addition, remote workers were the target population in this study; these individuals use the Internet daily, and some extent of generalizability is therefore guaranteed. Second, although there are several measurements of loneliness [27], in the present study, the presence of loneliness was assessed through a single question. However, this approach follows previous research that used a single item to measure loneliness [27]. Third, the causal relationship between remote work and the presence of loneliness is unknown because this was a cross-sectional study. Concerns have been raised about the existence of reverse causality in this relationship because, for example, certain workers might not choose to work remotely because they wish to avoid loneliness. However, during the COVID-19 pandemic, workers were not likely to be able to decide the frequency of telecommuting on their own, and the possibility of reverse causality is therefore probably low.

In conclusion, among remote workers in Japan, we found that support from co-workers and support from supervisors were strongly associated with loneliness, and frequency of remote work showed a moderate association with loneliness. These findings suggest that co-worker support and supervisor support may be crucial factors in preventing loneliness caused by working remotely. To prevent remote workers from feeling lonely and from developing mental health problems following loneliness, remote workers should engage in interaction with supervisors and co-workers using the ICT developed for this purpose.

## Key points

### What is already known about this subject

- There has been an increase in loneliness since the outbreak of COVID-19.
- Even before COVID-19, loneliness was a critical issue not only for older adults but also for the working generation.
- Telework can induce feelings of loneliness and isolation.

### What this study adds

- The frequency of telecommuting was moderately associated with loneliness.
- Support from co-workers and support from supervisors were strongly associated with feelings of loneliness.
- A high level of psychological job demands was associated with loneliness, whereas decision latitude was not associated with loneliness.

### What impact this may have on practice or policy

- Supervisors and co-workers may play an important role in preventing remote workers from feeling lonely and from developing mental health problems following loneliness.
- Decreasing psychological job demands may reduce the likelihood of loneliness for remote workers.

## Data Availability

No additional data are available.

## Acknowledgements

We thank the other members of the CORoNaWork Project: Yoshihisa Fujino (present chairperson of the study group), Hajime Ando, Hisashi Eguchi, Arisa Harada, Kyoko Kitagawa, Kosuke Mafune, Ryutaro Matsugaki, Koji Mori, Keiji Muramatsu, Masako Nagata, Ning Liu, Akira Ogami, Rie Tanaka, and Kei Tokutsu.

